# Development and Preclinical Validation of DiaBuddy™, a Point-of-Care Decision Support Tool for Children with Type 1 Diabetes: A Pilot Clinical Study

**DOI:** 10.64898/2026.03.10.26347874

**Authors:** Alapan Mahapatra, Sandeep Paimode, Sayan Banerjee, Prashant Shukla, Nimisha Sachan, Vibha Yadav, Anurag Bajpai

**Affiliations:** Department of Pediatric Endocrinology, Regency Center for Diabetes, Endocrinology & Research Kanpur; Department of Pediatric Endocrinology, Advanced Pediatric Center, Postgraduate Institute of Medical Education & Research, Chandigarh

**Keywords:** Decision Support, Diabetic Ketoacidosis, DiaBuddy, Insulin dose calculator, Sick Day Management, Type 1 Diabetes

## Abstract

**Background:** Optimal management of Type 1 diabetes (T1D) requires precise insulin dosing and sick-day decision-making; deficiencies in either predispose to hypoglycemia, suboptimal glycaemic control, and diabetic ketoacidosis. We developed DiaBuddy™, a guideline-aligned mobile decision support tool, and evaluated its accuracy versus expert guidance and its clinical impact in children with T1D.

**Methods:** The preclinical validation compared DiaBuddy and family recommendations (N = 37) for 20 insulin-dosing and 20 sick-day vignettes against a pediatric endocrinologist gold standard. The prospective single-arm pilot study evaluated its impact on HbA1c, CGM metrics, and quality of life in 20 children.

**Results:** In the preclinical study, DiaBuddy™ showed lower absolute relative deviation for basal (5.0 ± 6.7% vs 24.2 ± 25.9%), bolus (6.9 ± 10.9% vs 45.3 ± 48.8%), and combined doses (6.3 ± 9.3% vs 39.0 ± 37.8%; all P < 0.001) than families. DiaBuddy™ achieved ≥90% accuracy across all sick-day domains versus 27–70% for families; adherence would have prevented 94.5% of family errors including all hospitalization decision errors. In the clinical study, HbA1c declined from 9.18 ± 1.99% to 8.48 ± 1.44% (P = 0.049), PedsQL QOL score increased from 76.5 ± 8.6 to 89.1 ± 7.1 (+12.6 points, P < 0.001) with no change in CGM metrics. Application satisfaction was high (mean score 44.1 ± 4.1 out of 50) with 96% wishing to continue using it.

**Conclusions:** DiaBuddy delivers guideline-aligned guidance, substantially outperforming family decisions in preclinical vignette testing and showing preliminary signals of improvement in glycaemic control and quality of life in a small uncontrolled pilot study. The results support the conduct of larger randomised controlled trials.

## Introduction

Type 1 diabetes (T1D) is the most challenging chronic endocrine condition affecting children and adolescents worldwide, with rapidly rising global incidence [1]. Optimal day-to-day management requires frequent insulin dose adjustments based on pre-meal glucose concentrations, carbohydrate content, personalized parameters (insulin-to-carbohydrate ratio [ICR] and insulin sensitivity factor [ISF]), and the impact of intercurrent illness [2]. Failure to implement these predisposes children to recurrent glycemic excursions, suboptimal control, and life-threatening diabetic ketoacidosis (DKA) [3].

Consistent carbohydrate counting improves glycemic control and reduces postprandial excursions [4], but real-world implementation is undermined by limited access to food composition data and attrition of skills over time [5, 6]. Inappropriate sick-day management is the leading cause of DKA in children with T1D [7, 8], yet routine adherence to evidence-based sick-day protocols remains poor. There is therefore a compelling need for accessible, point-of-care decision support that empowers families to deliver rational insulin dosing and implement sick-day protocols reliably between clinic visits.

Bolus calculators and digital carbohydrate-counting programs have been shown to reduce postprandial glycemic excursions and hypoglycemia [9–13], but the majority address only meal-time insulin management without integrating sick-day logic. The present author group has developed and validated mobile decision-support applications for short stature, severe DKA, bone-age assessment, and pediatric diabetes interpretation [14–17]. Building on this experience, they developed DiaBuddy, a comprehensive mobile application for pediatric T1D management that integrates ISPAD guideline–aligned algorithms for basal titration, bolus dosing, and sick-day care alongside modules for meal planning, activity guidance, glucose logging, and patient education.

This study reports the preclinical vignette-based validation of DiaBuddy against a paediatric endocrinologist gold standard and pilot clinical evaluation of its impact on glycemic control and health-related quality of life in children with T1D.

## Methodology

### DiaBuddy™ Application

DiaBuddy™ is a comprehensive point-of-care decision support application for pediatric T1D with seven integrated modules: (1) Insulin Wizard (basal titration, bolus dosing, and correction doses), (2) Sick-Day Guide (stepwise intercurrent illness management), (3) Sugar Tracker (glucose log with customizable alerts), (4) EatRite (personalized meal planning with carbohydrate quantification), (5) PlayRite (physical activity guidance and tracking), (6) SleepRite (sleep hygiene monitoring), and (7) Knowledge Base (patient and family educational resources across T1D domains). This study evaluates the Insulin Wizard and the Sick-Day Guide.

At enrolment, the clinical team enters the child’s honeymoon status, insulin regimen (basal-bolus MDI, split-mixed, CSII, or premix), insulin type, doses, and timing. The application computes the ICR (500/TDD) and ISF (1,800/TDD for rapid-acting; 1,500/TDD for regular insulin). Bolus doses integrate pre-meal glucose, meal carbohydrate content from CarbCal (a proprietary nutrition database of 10,245 foods), and a default meal target of 120 mg/dL (modifiable by the clinician): Bolus dose = [Carbohydrate (g) / ICR] + [(BG − 120) / ISF]. Basal dose titration follows fasting glucose–linked multipliers (×0.8 for <70 mg/dL through ×1.2 for >200 mg/dL). Insulin-on-board is estimated via linear decay at 20% per hour over 5 hours, and correction doses are computed as [(BG − 140) / ISF] − IOB for post-meal hyperglycaemia exceeding 300 mg/dL. All algorithms are anchored to ISPAD 2022 guidelines [2, 18–20].

The Sick-Day Guide requests clinical features (vomiting, abdominal pain, rapid breathing), capillary blood glucose, and blood or urine ketone values. The algorithm generates ISPAD-aligned recommendations on hospital admission, TDD modification, supplemental rapid-acting insulin, and oral fluid type and volume (6 mL/kg/hour, with sugar-free fluids when glucose ≥250 mg/dL). Hospital referral is triggered by clinical signs of DKA or blood ketones >3.0 mmol/L or urine ketones 3–4+.

### Phase 1: Preclinical Vignette-Based Validation

Thirty-seven families of children with T1D completed 20 standardized insulin-dosing (12 bolus, 8 basal) and 20 sick-day vignettes, blinded to DiaBuddy outputs. Vignettes were drafted by two pediatric endocrinologists and independently reviewed for guideline concordance. A pediatric endocrinologist affiliated with a different institution from the primary research team and with no financial interest in DiaBuddy, was engaged as the independent gold standard. He was blinded to the family and application outputs throughout the adjudication process and provided the reference answer for each scenario.

### Phase 2: Pilot Clinical Study

A prospective, single-arm study enrolled 25 children and adolescents with T1D (diabetes duration >1 year, age 5–18 years) attending our pediatric endocrinology clinic. The Phase 2 cohort was independent of the Phase 1 vignette study; no participant was enrolled in both phases.

Two participants with incomplete follow-up data and three who withdrew voluntarily were excluded, leaving 20 participants for analysis. At baseline, participants underwent measurement of hemoglobin A1c (HbA1c), 15 days of intermittently scanned continuous glucose monitoring with FreeStyle Libre 2 (Abbott), and assessment of quality of life using the PedsQL questionnaire [21]. Families then received structured training on the use of DiaBuddy and were provided unrestricted access to the application for home use. After three months of DiaBuddy use, HbA1c, intermittent continuous glucose monitoring metrics and quality-of-life scores were reassessed. Satisfaction with application use was evaluated using a bespoke 10-item Likert-type rating scale (maximum 50 points), developed a priori by the study team to assess usability, perceived clinical utility, and overall satisfaction. Each item was scored 1–5, with higher scores indicating greater satisfaction. Families were also asked whether they wished to continue using the application after study completion.

### Statistical analysis

Continuous variables were summarized as mean ± standard deviation (SD) and categorical variables as counts and percentages. For the preclinical study accuracy of insulin dose recommendations was quantified using absolute relative deviation (ARD), defined as |recommendation − gold standard| / gold standard × 100%. Agreement between DiaBuddy and the expert gold standard was evaluated using Bland–Altman analysis. Sick-day accuracy was evaluated by domain: hospitalization decision, TDD modification (accurate if within 10%), fluid type, and supplemental rapid-acting insulin dose (accurate if within 10%). Domain-specific weights were pre-specified (hospitalization: 4; supplemental rapid-acting insulin: 3; TDD modification: 2; fluid type: 1). Weighted net benefit and prevention index were calculated.

Baseline and post-intervention outcomes were compared using paired t-tests (normally distributed) or Wilcoxon signed-rank tests (non-normal distribution). Effect sizes for within-subject changes were calculated using Cohen’s dz for paired samples with changes presented as mean and 95% confidence intervals (CI). Clinically meaningful improvement in glycemic control was predefined as a reduction in HbA1c ≥0.5%. All statistical tests were two-sided, and p values <0.05 were considered statistically significant. The frequency of hypoglycemia episodes before and during DiaBuddy use was compared using the χ² test. Analyses were performed in SPSS v23 (Windows). The protocols were approved by the Institutional Ethics Committee of our hospital, and written informed consent/assent was obtained prior to participation. The study protocol conforms to the Declaration of Helsinki.

## Results

### Phase 1: Preclinical Validation

Thirty-seven children with T1D (19 boys; mean age 15.2 ± 0.7 years; diabetes duration 6.1 ± 0.6 years) participated. All participants were on multiple daily injections using analog insulins. Mean HbA1c was 10.2 ± 2.4%. Mean annual severe hypoglycemia and DKA events were 2.3 ± 1.0 and 0.3 ± 0.6 respectively. Self-monitoring of blood glucose was performed 2.4 ± 0.5 times/day; 8 participants (21.7%) reported regular carbohydrate counting.

DiaBuddy showed lower ARD compared with families for basal (5.0 ± 6.7% vs 24.2 ± 25.9%), bolus (6.9 ± 10.9% vs 45.3 ± 48.8%), and combined doses (6.3 ± 9.3% vs 39.0 ± 37.8%; all P < 0.001; Table 1). Of 730 family responses, 308 (42.1%) were below and 107 (14.6%) above the ±20% error margin; the corresponding numbers for DiaBuddy™ were one each (5%). Bland-Altman plots demonstrated narrower limits of agreement for DiaBuddy relative to the gold standard than for family recommendations (Figure 2). Family bolus dose bias was −2.0 ± 9.2 units versus −0.5 ± 2.0 units for DiaBuddy (P < 0.001). Participants practicing regular carbohydrate counting had lower bolus ARD (28.7 ± 5.4% vs 49.9 ± 54.3%, P = 0.04) and fewer annual DKA episodes (0 vs 0.4 ± 0.6; P = 0.002) than those not counting carbohydrates.

**Figure 1.**
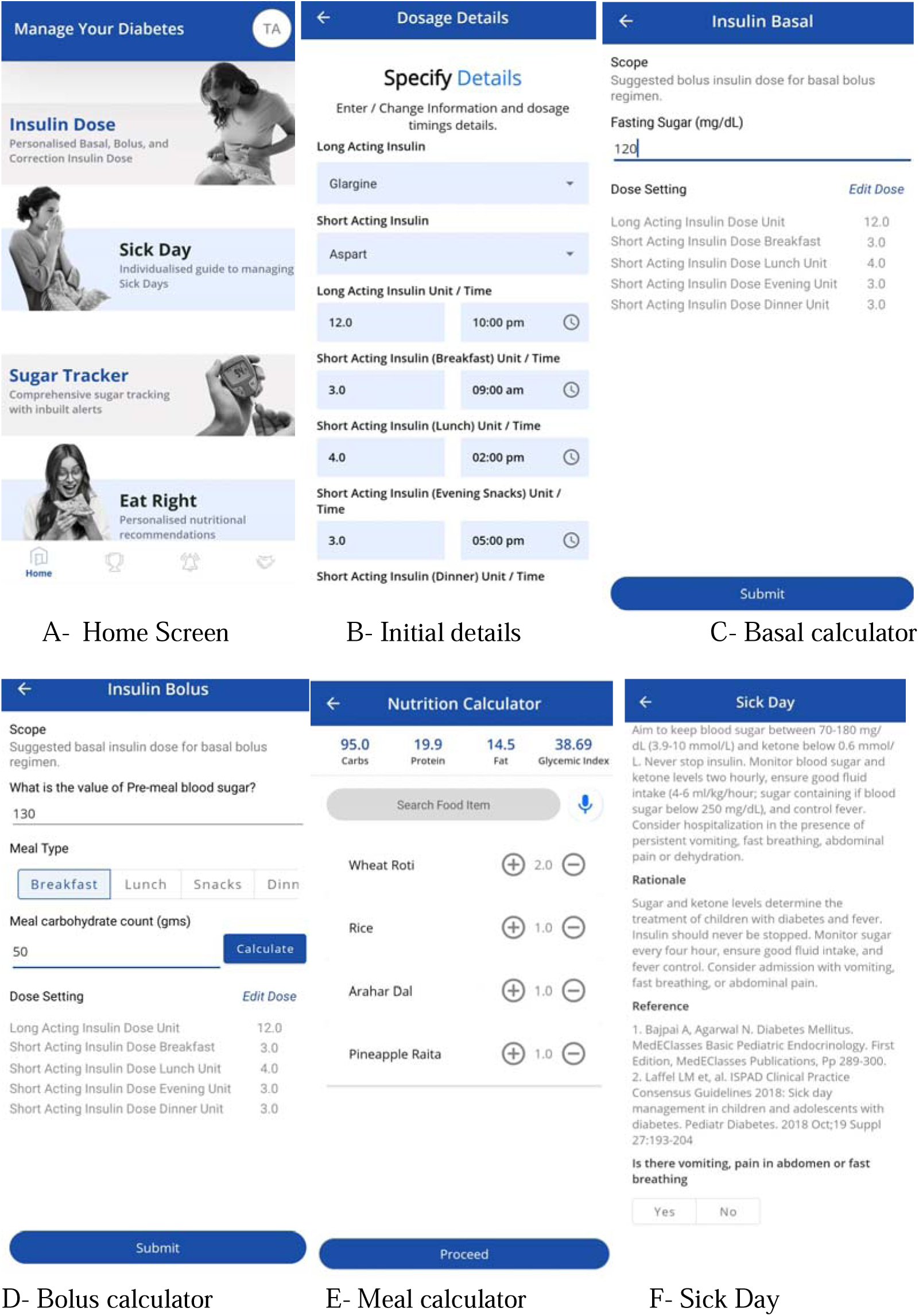
Workflow of the mobile application 12

**Figure 2.**
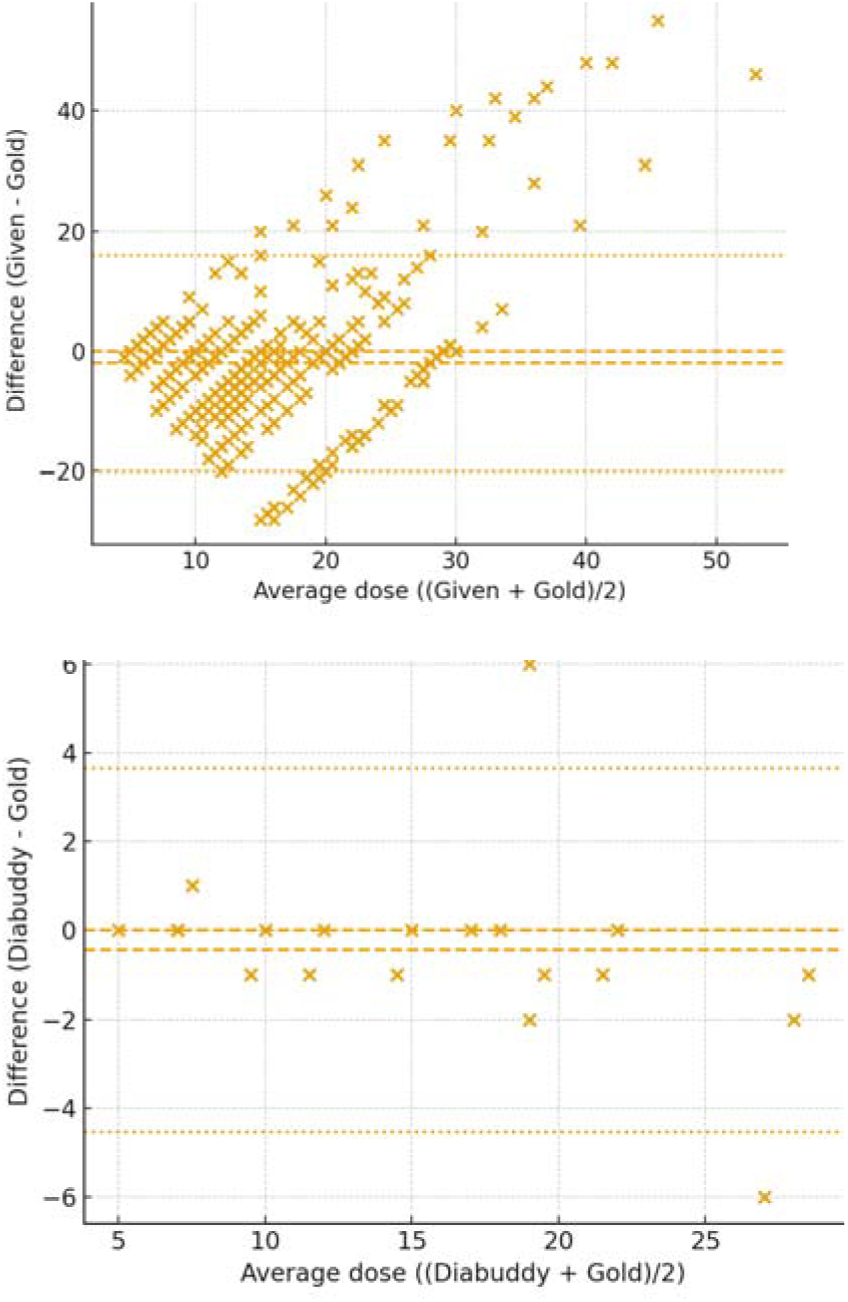
Bland Altman Plot for difference between participants and Diabuddy compared to gold standard

**Figure 3.**
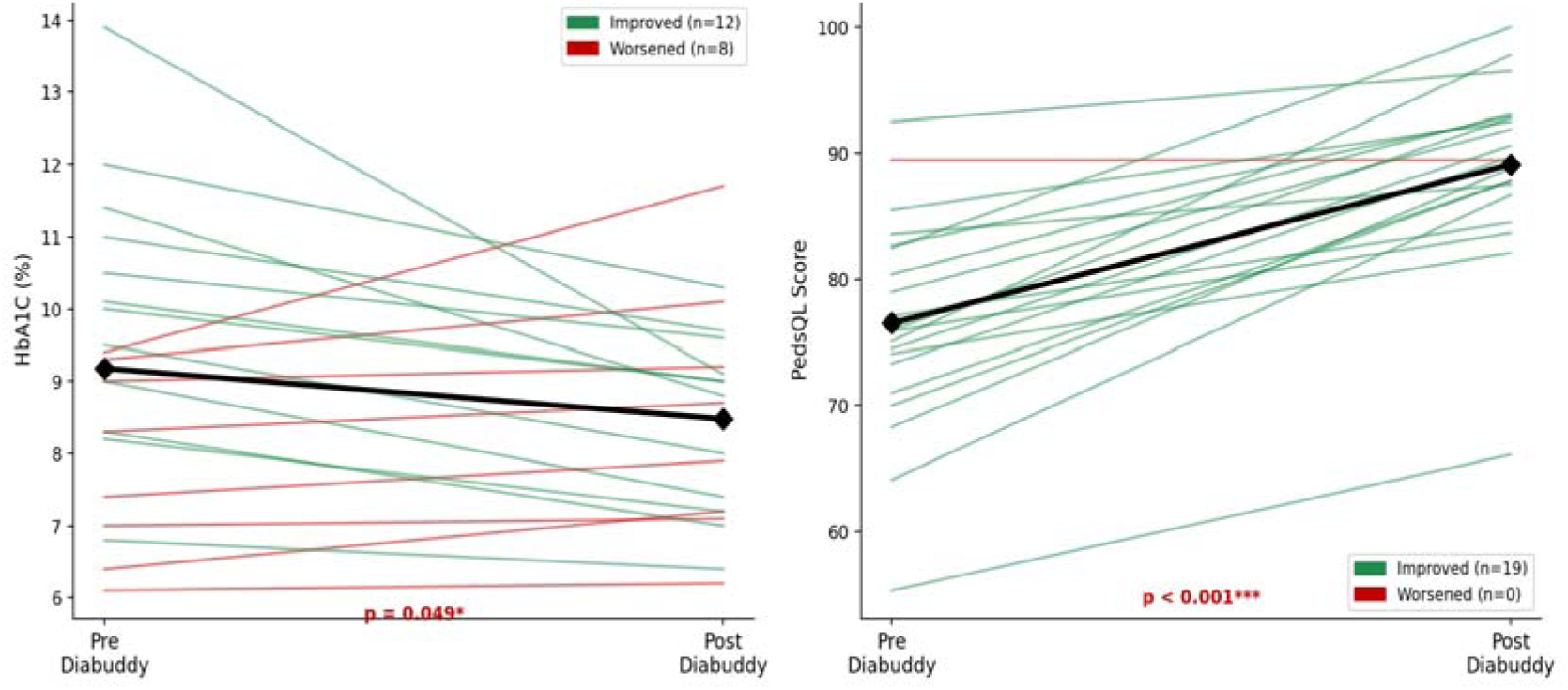
Individual Hemoglobin A1c and PedsQL quality-of-life trajectories pre- and post-DiaBuddy™. Bold black line = group mean ± SE. P < 0.001.

**Table 1.** Absolute Relative Deviation (%) for basal, bolus, and combined insulin doses: Family vs DiaBuddy compared to gold standard (N = 37, 20 vignettes each)

| Category | Family | DiaBuddy | P-value |
| --- | --- | --- | --- |
| Basal insulin ARD (%) | 24.2 ± 25.9 | 5.0 ± 6.7 | < 0.001 |
| Bolus insulin ARD (%) | 45.3 ± 48.8 | 6.9 ± 10.9 | < 0.001 |
| Combined dose ARD (%) | 39.0 ± 37.8 | 6.3 ± 9.3 | < 0.001 |
| Doses within ±20% of target (%) | 43.3% | 95.0% | < 0.001 |
| Bolus dose bias (units) | -2.0 ± 9.2 | -0.5 ± 2.0 | < 0.001 |

ARD = absolute relative deviation. Doses within ±20% margin considered clinically acceptable. P-values from Welch's t-test.

DiaBuddy™ achieved ≥90% accuracy across all sick-day management domains, compared with 27–70% for families (Table 2). Family errors across 2,960 decision-circumstances totaled 1,532 (51.8%), including 223/740 (30.2%) hospitalization decisions, 547/740 (73.9%) TDD modifications, 500/740 (67.5%) supplemental rapid-acting insulin doses, and 262/740 (35.4%) fluid recommendations. Adherence to DiaBuddy guidance would have prevented 94.5% of these errors, including all family errors in hospitalization decisions. The weighted net benefit was 3,437 and the weighted prevention index was 0.950.

**Table 2.** Sick-day management accuracy, error prevention, and inter-rater agreement (N = 37, 20 vignettes each)

| **Decision Domain (Weight)** | **DiaBuddy**<br>**Accuracy** | **Family**<br>**Accuracy** | **Error**<br>**Prevention** |
| --- | --- | --- | --- |
| Hospitalization decision (4) | 100% | 70% | 100% |
| TDD modification (2) | 90% | 27% | 90.4% |
| Fluid type/volume (1) | 95% | 65% | 97.7% |
| Supplemental rapid-acting<br>insulin (3) | 95% | 33% | 94.8% |
| Overall (all domains) | ≥90% | 27–70% | 94.5% |

TDD = total daily dose. Clinical priority weights are shown in parentheses.
Prevention = proportion of family errors corrected by DiaBuddy™ adherence.

**Table 3.**
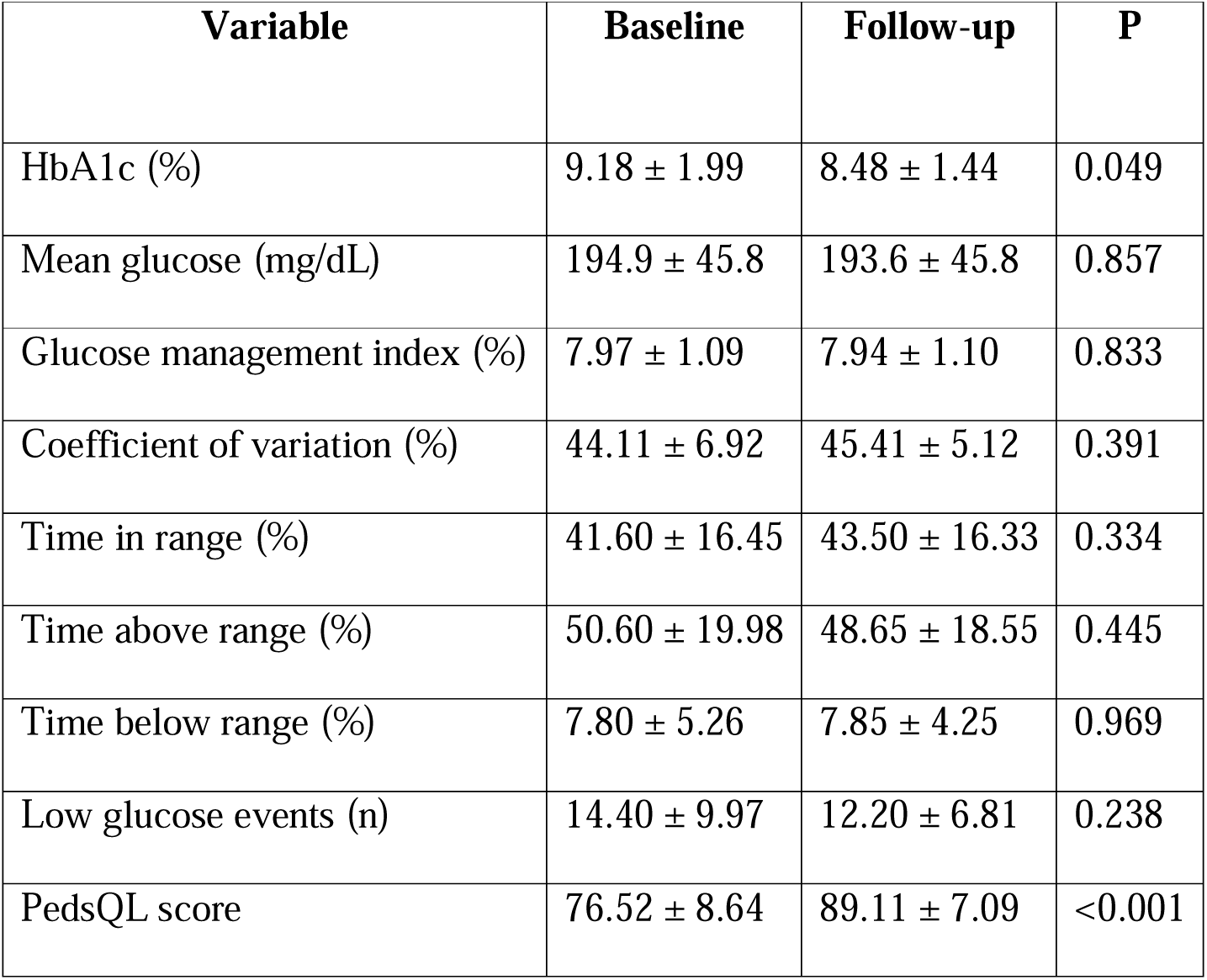
Changes in Clinical and CGM Parameters After Three Months of DiaBuddy Use (N = 20)

### Phase 2: Pilot Clinical Study

Pilot clinical validation was conducted in 25 children and adolescents with type 1 diabetes (13 boys). Twenty children (10 boys; mean age 12.47 ± 4.12 years; range 5.5–18 years) were included in the analysis; two with incomplete follow-up data and three who opted out of the study were excluded. Excluded participants were older (16.16 ± 2.43 vs 12.52 ± 4.19 years; p ≈ 0.024) and had similar baseline HbA1c (11.04 ± 2.64 vs 9.18 ± 1.99, p > 0.20), and gender distribution (60% versus 50% boys; p = 0.638) compared to those who completed the study.

After three months of DiaBuddy use, mean HbA1c decreased from 9.18 ± 1.99% at baseline to 8.48 ± 1.44% at follow-up, corresponding to a mean reduction of −0.70 ± 1.49% (95% CI −1.40 to −0.003; p = 0.049; Cohen’s dz = 0.47). HbA1c improved in 12 of 20 participants (60%), while 11 (55%) achieved a clinically meaningful reduction of at least 0.5%, and 9 (45%) had reductions of ≥1.0%. Mean blood glucose, glucose management indicator (GMI), coefficient of variation, time in range (TIR), time above range (TAR), and time below range (TBR) did not change significantly over the three-month follow-up period.

Diabetes-related quality of life improved following DiaBuddy with increased total PedsQL score from 76.52 ± 8.64 to 89.11 ± 7.09 with a mean improvement of +12.60 ± 6.48 points (95% CI +9.56 to +15.63; p < 0.001; Cohen’s dz = 1.94). Quality-of-life scores improved in 19 of 20 participants (95%), with 17 participants (85%) demonstrating clinically meaningful improvement.

The tool was used on a 66.4 ± 18.7% of days over the study period (range 34.4–95.6%); no significant association was observed between usage frequency of and HbA1c change (rs = +0.14; p > 0.20). Application satisfaction was high, with a mean satisfaction score of 44.1 ± 4.1 out of a maximum possible score of 50, and 96% of participants indicating that they wished to continue using the application after study completion.

## Discussion

This study provides evidence supporting the potential utility of DiaBuddy in implementing guideline-aligned insulin dosing and sick-day management for children with type 1 diabetes. In the vignette-based validation phase, DiaBuddy demonstrated greater accuracy than family-derived decisions while in the pilot clinical evaluation, three months of its use was associated with clinically meaningful reduction in HbA1c and improvements in diabetes-related quality of life.

The disparity in sick-day management accuracy between DiaBuddy and family recommendations represents a particularly important observation, given that inappropriate sick-day management is the leading cause of DKA in children with T1D [7, 8]. DiaBuddy achieved ≥90% accuracy across all decision domains compared with 27–70% for families, and adherence to its guidance would have prevented all family errors in hospitalization decisions the domain carrying the greatest clinical consequence. These findings align with and extend prior evidence on the value of point-of-care decision support in T1D [9–13], while uniquely addressing sick-day care as an integrated algorithmic module.

The disparity between family and DiaBuddy insulin-dosing accuracy is consistent with the literature documenting that without decision support, families struggle to accurately implement bolus calculations in real-world conditions [9, 12]. Family ARD for bolus dosing was nearly seven times higher than DiaBuddy (45.3% vs 6.9%), and fewer than half of family recommendations fell within the clinically acceptable ±20% margin compared with 95% for DiaBuddy. The correlation between carbohydrate-counting practice and lower ARD and fewer DKA events reinforces the importance of nutritional literacy in T1D management [4], and positions DiaBuddy’s CarbCal tool as an asset in resource-limited settings.

The HbA1c reduction of −0.70 percentage points over three months is directionally consistent with meta-analytic estimates of 0.3–0.9% for smartphone-based interventions in pediatric T1D [9]. More than half of participants achieved a clinically meaningful reduction of at least 0.5%. These findings should be interpreted cautiously: the single-arm uncontrolled design cannot distinguish DiaBuddy-specific effects from regression to the mean, structured training effects, or increased clinical contact. Lack of significant change in CGM metrics likely reflects the study’s short duration, modest sample size, and the substantial within-person CGM variability. Longer-term follow-up with a larger sample is required to detect statistically significant CGM metric improvements.

The improvement in quality-of-life score is the most striking finding of the clinical phase, with a mean PedsQL gain of +12.6 points and a large effect size (Cohen’s dz = 1.94). This suggests that DiaBuddy may meaningfully reduce the day-to-day psychological burden of diabetes management for families, consistent with evidence linking decision support tools to reduced diabetes distress and improved treatment adherence [2]. The high satisfaction score (44.1/50) and the near-universal desire to continue using the application (96%) further attest to the platform’s acceptability.

Strengths of this study include the use of an independent blinded expert as the gold standard, ISPAD 2022 guideline alignment, and a structured two-phase design that bridges preclinical and clinical evaluation. Several limitations must be acknowledged. The vignette-based design of the preclinical phase simulates, but does not replicate, real-world family decision-making under time pressure and emotional stress; families may perform differently in live clinical encounters. All Phase 1 participants were managed with basal-bolus MDI; algorithm performance for split-mixed, CSII, and premix regimens was not independently validated. The clinical phase lacked a control arm, precluding causal attribution of observed improvements to DiaBuddy specifically; regression to the mean, Hawthorne effect, and the effect of structured training independent of the application are alternative explanations that cannot be excluded. The three-month follow-up was insufficient to detect changes in CGM metrics, and the sample was not powered for these secondary endpoints. The age difference between excluded and analyzed participants suggests that findings may not generalize to older adolescents and young adults with T1D.

To conclude, DiaBuddy offers a scalable, low-cost digital solution for delivering guideline-aligned care to children with T1D, particularly in settings where structured diabetes education and specialist contact are infrequent. Future research should include multicenter randomized controlled trials with longer follow-up, and health-economic evaluation in low-and-middle-income settings where the unmet need is greatest.

## Data Availability

All data produced in the present study are available upon reasonable request to the authors

## ACKNOWLEDGEMENTS

The authors thank the children and families who participated in both study phases. We acknowledge the contribution of the nursing and diabetes educator teams at the Regency Center for Diabetes, Endocrinology & Research, Kanpur.

## Funding

This study received no specific grant from any funding agency in the public, commercial, or not-for-profit sectors.

## Conflicts of interest

A.B. is a co-developer of DiaBuddy™. To mitigate potential bias, A.B. was not involved in vignette construction, family data collection, or the selection and engagement of the independent expert gold standard. S.B., affiliated with a different institution from the primary research team and with no financial interest in DiaBuddy, was the independent gold standard.

## Ethics statement

This study was performed in accordance with the Declaration of Helsinki. This human study was approved by Regency Hospital Limited IEC-approval: 16084, 160152, 160151. All parents, guardians or next of kin provided written informed consent for the minors to participate in this study.

## Author contributions

A.M., S.P., and A.B. conceptualized and designed the study. A.M., S.P., P.S., N.S., and V.Y. collected data. S.B. was the independent gold standard. A.M. and A.B. wrote the manuscript. A.B. will act as the guarantor of the study. All authors reviewed and approved the final version.

## Data Availability

On request

## Notes

### Clinical Trial

NCT07470905

### Funding Statement

This study did not receive any funding

### Author Declarations

This study was performed in accordance with the Declaration of Helsinki. This human study was approved by Regency Hospital Limited's Institutional Ethics Committee with approval numbers 16084, 160152, and 160151. All parents, guardians, or next of kin provided written informed consent for the minors to participate in this study.

